# Enzyme immunoassay for SARS-CoV-2 antibodies in dried blood spot samples: A minimally-invasive approach to facilitate community- and population-based screening

**DOI:** 10.1101/2020.04.28.20081844

**Authors:** Thomas W. McDade, Elizabeth M. McNally, Richard D’Aquila, Brian Mustanski, Aaron Miller, Lauren A. Vaught, Nina L. Reiser, Elena Bogdanovic, Aaron S. Zelikovich, Alexis R. Demonbreun

## Abstract

**Background:** Serological testing for SARS-CoV-2 IgG antibodies is needed to document the community prevalence and distribution of the virus, particularly since many individuals have mild symptoms and cannot access molecular diagnostic testing of naso-pharyngeal swabs. However, the requirement for serum/plasma limits serological testing to clinical settings where it is feasible to collect and process venous blood. To address this problem we developed a serological test for SARS-CoV-2 IgG antibodies that requires only a single drop of capillary whole blood, collected from a simple finger prick and dried on filter paper (dried blood spot, DBS).

**Methods:** Enzyme linked immunosorbent assay (ELISA) was optimized to detect SARS-CoV-2 IgG antibodies against the receptor-binding domain (RBD) of the spike protein. DBS samples were eluted overnight and transferred to a 96-well plate coated with antigen, and anti-human IgG-HRP was used to generate signal in proportion to bound antibody. DBS samples spiked with anti-SARS IgG antibody, and samples from known positive and negative cases, were compared to evaluate assay performance.

**Results:** Analysis of samples with known concentrations of anti-SARS IgG produced the expected pattern of dose-response. Optical density (OD) values were significantly elevated for known positive cases in comparison with samples from unexposed individuals.

**Discussion:** DBS ELISA provides a minimally-invasive alternative to venous blood collection that combines the convenience of sample collection in the home or non-clinical setting with the quantitation of ELISA in the lab. Serological testing for SARS-CoV-2 IgG antibodies in DBS samples should facilitate research across a wide range of community- and population-based settings on seroprevalence, predictors and duration of antibody responses, as well as correlates of protection from reinfection, each of which is critically important for pandemic control.

## Introduction

Severe acute respiratory syndrome coronavirus 2 (SARS-CoV-2)—the virus that causes coronavirus disease 2019 (COVID-19)—is a global pandemic with no vaccine or established pharmacological treatment ^1^. Testing for infection is key to identifying cases and for tracing community spread of the virus, which is critical for informing efforts aimed at mitigation. Provider-collected nucleic acid-based tests of naso-pharyngeal (NP) swabs are being used worldwide to detect virus during the acute stage of infection, but shortages of personal protective equipment (PPE), NP swabs, transport media, and testing reagents in the US have limited application to the most serious cases, inhibiting efforts to identify and trace the actual prevalence and spread of the virus in the community. In addition, viral RNA production in the naso-pharynx following infection may be transient, intermittent and/or subject to sampling variability, contributing to false negative results.

Serological testing is a complementary approach that detects the presence of antibodies against SARS-CoV-2 in blood samples from exposed individuals^2^. Antibodies specific to viral proteins will emerge 3-10 days after infection, with IgM appearing first (transiently) and IgG becoming detectable approximately 14 days after infection^3^. IgG antibodies remain detectable in blood after the symptoms of infection resolve, and can therefore be used to identify "cases” months after recovery (based on limited duration of longitudinal study to date)^4,5^.

Serological testing for SARS-CoV-2 offers the following advantages: 1) It provides data to inform estimates of the seroprevalence of infection and case fatality rates, which can only be estimated by testing individuals who are infected but show no symptoms, or only mild symptoms, of disease; 2) It can be used to illuminate the geographic spread of the virus, and to identify subgroups of individuals more susceptible to infection; and 3) It can enable research into correlates of protection from SARS-CoV-2 reinfection and their duration. Much of our knowledge of COVID-19 has derived from the most severe hospitalized and even lethal cases, with far less information coming from milder cases in the community. Additional research is needed before serological testing can be used to answer critical questions such as who is safe from reacquisition of COVID-19 or how to roll out vaccine access to those still susceptible.

SARS-CoV-2 IgM and IgG antibodies can be quantified with enzyme linked immunosorbent assay (ELISA), and protocols for serum or plasma samples are now established^6^. However, the use of serum/plasma limits wider application in non-clinical settings due to the costs and logistical constraints associated with collecting, processing, and transporting venous blood. Deploying phlebotomists for large scale testing would also consume scarce PPE. Furthermore, when sheltering-in-place is required or encouraged, people may be less able or willing to leave home and samples drawn for serology studies that rely on clinic-based blood collection are likely to be biased.

There is an alternative in finger stick dried blood spot (DBS) sampling^7^. With DBS, a lancet is used to prick the finger, and up to five drops of blood are placed on filter paper, where it dries. DBS sampling has served as the foundation for nation-wide newborn screening programs since the 1960s, and it is increasingly applied as a minimally-invasive alternative to venipuncture in community- and population-based health research, including several applications in infectious disease epidemiology^8–10^. Advantages of DBS sampling include low cost, potential for self-sampling, ease of shipment (samples can be returned in the mail with no cold chain or special handling required), and reduced biohazard risk in transit and in the lab (the CDC and US Postal Service consider DBS specimens nonregulated, exempt materials)^11^. It is important to emphasize that DBS sampling is distinct from point-of-care lateral flow (LF) testing, which also uses finger stick blood. Recent analyses have raised significant concerns regarding the accuracy of LF tests for SARS-CoV-2 IgG antibodies^12^. Our approach combines the convenience of finger stick blood collection with the analytic rigor that can be applied in the laboratory.

Below we describe two ELISA protocols for measuring SARS-CoV-2 IgG antibodies in DBS samples. The first is an adaptation of a widely disseminated serum protocol^6^. The second is a more streamlined protocol based on a previously validated DBS ELISA^13^. Both protocols focus on IgG because it remains detectable long after infection, although the protocols can be easily adapted to measure IgM. The assays target the receptor-binding domain (RBD) of the spike protein, a large glycoprotein on the viral surface that mediates attachment to host cells and facilitates viral entry. The RBD is very immunogenic: It is the target of neutralizing antibodies, and prior work has demonstrated that it is an effective capture antigen in ELISA^6,14,15^. We describe our protocols as well as results from the analysis of confirmed positive and negative samples to demonstrate the validity of the DBS assays.

## Methods

The assay principle is as follows: DBS samples are eluted overnight, and eluate is transferred to microtiter wells, which were previously coated with SARS-CoV-2 RBD antigen. Goat anti-human IgG-HRP binds to the RBD-antibody complexes in each well, and color forms with the addition of chromogenic substrate. The absorbance of the solution is read at 490nm.

### Reagents and supplies

#### Filter papers

Whatman #903 (Cytiva #10534612). These cards are approved by the FDA as Class II medical devices for diagnostic applications, and the CDC implements an independent quality control analysis of each card lot to confirm thickness, flow-rate, absorbency, and purity.

#### Hole punch

There are multiple options for punching out uniform discs of whole blood from DBS samples. We used a semi-automatic pneumatic system with 5mm punch (Analytical Sales and Services #327500, Flanders, NJ). Manual punches are also readily available that produce 3/16 inch discs (4.8mm) (e.g., McGill #MCG53600C). A 5 mm punch will contain approximately 5uL of serum^9,16^.

#### Microtiter plates

For elution of DBS samples: Corning 96 Well TC-Treated Microplates (Sigma #CLS3599). For coating with antigen: Immulon 4 HBX polystyrene 96-well flat bottomed, high-binding plates (ThermoFisher Scientific #3855).

#### Coating antigen

The plasmid for the receptor binding domain (RBD) of the spike (S) glycoprotein gene from SARS-CoV-2, Wuhan-Hu-1 (GenBank: MN908947) was generously provided by Dr. Florian Krammer (Mt. Sinai Medical School, NY) and is described previously^6^. The construct was made by fusing the N-terminal S protein signal sequence to the spike RBD (amino acids 419 to 541) with a C-terminal hexa-histidine tag. The sequence was codon optimized for mammalian expression and subcloned into the pCAGGS mammalian expression vector. Recombinant RBD protein was generated by Evotec using standard methods (Princeton, New Jersey). Briefly, EXPI293 cells were transfected with purified, endotoxin free plasmid DNA. Four L of media was purified using IMAC chromatography 72 hours post transfection. The final recovery of purified recombinant RBD protein was ~195mg in PBS with an endotoxin level at ~ 1EU/mg, with a purity >95%.

#### Detection antibody

Anti-Human IgG (Fab specific)-Peroxidase antibody produced in goat (Sigma-Aldrich #A0293).

#### Positive control

IgG antibody to SARS-CoV S protein (CR3022 antibody, Creative Biolabs #MRO-1214LC).

#### Coating buffer

Phosphate buffered saline (1x PBS; Fisher BP24384 or equivalent)

#### Chromogenic substrate

SIGMA*FAST*™ OPD (Sigma-Aldrich #P9187). One silver and one gold tablet per 2 plates in 20 mL dH2O.

#### Stop solution

3.0M Hydrochloric acid (HCl).

The above reagents and supplies are used in both protocols. The following reagents differ:

*Protocol 1 wash buffer* PBS, 0.1% Tween 20 (PBS-T; Fisher BP337–500).
*Protocol 1 blocking solution* 0.1% PBS-T, 3% milk (w/v; American Bio AB10109-01000).
*Protocol 1 antibody diluent* 0.1% PBS-T, 1% milk (w/v).
*Protocol 2 assay buffer (for blocking, washing, and antibody dilution)* 0.01 M phosphate buffer, 0.5 M NaCl, 0.1% Tween 20.

### Preparation of materials

DBS calibrators are prepared as follows: 1) Dilute CR3022 antibody to desired concentrations in PBS; 2) Collect EDTA whole blood from a negative donor; 3) Add CR3022 dilutions to aliquots of whole blood, mixing gently to avoid hemolysis; and 4) Transfer with a pipet (65 μL/drop) to labeled #903 cards. Dry overnight at room temperature and store at −23°C. It is important to minimize the volume of CR3022 dilution that is added to whole blood, as large volumes (>5% of total volume) will lower hematocrit and adversely affect how the sample is absorbed on the filter paper.

### Protocol 1

The 96-well plate is coated with antigen as follows: Dilute RBD to 2 μg/mL in PBS, add 100 μL to each well, seal, and incubate overnight at 4°C. DBS samples are eluted overnight. Punch out one 5.0 mm disc of each DBS calibrator, control, and sample, and place in elution plate. Add 250 μL PBS, seal the plate, and elute overnight at 4°C.

The following day, remove plate and wash 4x with 300 μL PBS-T (BioTek ELx50). Add 200 μL per well 0.1% PBS-T, 3% milk. Block covered at room temperature for two hours. Aspirate.

At one hour, remove DBS samples from refrigerator and rotate for at 300 rpm (Heidolph Titramax 101). After removing blocking solution, transfer 100 μL eluate from DBS calibrators, controls, and samples to the coated 96-well plate. Cover the plate and incubate at room temperature for two hours. Wash the plate four times with PBS-T.

Prepare a dilution (1:3000) of detection antibody in 0.1% PBS-T, 1% milk. Add 100 μL working antibody solution to each well. Cover and incubate at room temperature for 60 minutes. Wash four times as before.

Prepare chromogenic substrate by dissolving one set of tablets in 20mL dH2O. Do not add silver tablet until ready to use. Add 100 μL chromogenic substrate to each well. Cover the plate, protect from the light, and incubate for 10 minutes at room temperature. Add 50 μL 3M HCl stop solution to each well. Incubate 5 minutes at room temperature. Read the absorbance (optical density, OD) at 490 nm (BioTek ELx808).

### Protocol 2

The full protocol is described, although it is very similar to Protocol 1. The key differences are the use of a single buffer for elution, blocking, and washing, and a shorter assay duration (3 hours 15 minutes total incubation time vs. 5 hours 15 minutes for Protocol 1).

The 96-well plate is coated with antigen as follows: Dilute RBD to 2 μg/mL in PBS, add 100 μL to each well, seal, and incubate overnight at 4°C. DBS samples are eluted overnight. Punch out one 5.0 mm disc of each DBS calibrator, control, and sample, and place in elution plate. Add 250 μL assay buffer, seal the plate, and elute overnight at 4°C.

The following day, remove DBS samples from refrigerator and rotate for one hour at 300 rpm (Heidolph Titramax 101). At 30 minutes, remove the coated 96-well plate and wash four times with Assay Buffer (BioTek ELx50). After final wash leave 300 μL Assay Buffer in each well. Incubate 30 minutes at room temperature, then aspirate.

Transfer 100 μL eluate from DBS calibrators, controls, and samples to the coated 96-well plate. Cover the plate and rotate at 250 rpm, room temperature, for 90 minutes. Wash the wells four times with Assay Buffer.

Prepare a dilution (1:3000) of detection antibody in Assay Buffer. Add 100 μL working antibody solution to each well. Cover and rotate at 250 rpm at room temperature for 60 minutes. Wash four times as before.

Prepare chromogenic substrate by dissolving one set of tablets in 20mL dH2O. Do not add silver tablet until ready to use. Add 100 μL chromogenic substrate to each well. Cover the plate, protect from the light, and incubate for 10 minutes at room temperature. Add 50 μL 3M HCl stop solution to each well. Incubate 5 minutes at room temperature. Read the absorbance (optical density, OD) at 490 nm (BioTek ELx808).

### Validation

We used a dilution series of CR3022, an IgG to SARS-CoV known to react with SARS-CoV-2 RBD, to confirm antibody capture and optimize the assay. EDTA venous whole blood from a negative donor was collected and spiked with small volumes of CR3022 in PBS at the following concentrations (μg/mL; relative to whole blood volume): 25.00, 12.50, 6.25, 3.13, 1.56, 0.78, 0.39, 0.20, 0.01, 0.00. Spiked whole blood was gently mixed and transferred to filter paper in 65 μL drops, allowed to dry four hours, then stored in gas impermeable bags with desiccant at - 23°C. The day before the assay, wells were coated with 2.0 μg/mL RBD in PBS, as described above. A separate set of wells was also coated with PBS (0.0 μg/mL RBD). Two sets of CR3022 DBS materials were punched out, eluted in 250 μL PBS or assay buffer, and transferred to the 2.0 and 0.0 μg/mL conditions the following day. The protocol was implemented as described above, and OD readings were compared across the conditions.

Finger stick DBS samples were collected from nine individuals with confirmed cases of COVID-19 based on PCR positive tests for the presence of SARS-CoV-2 in naso-pharyngeal swabs. Mean number of days between PCR test and DBS collection was 26.7 (range: 25–35). DBS samples from five individuals presumed to be negative based on the pre-pandemic date of sample collection (2018) were analyzed on the same plate. All samples were analyzed in duplicate, with the average result reported. All samples were de-identified and all research activities were implemented under protocols approved by the institutional review board at Northwestern University (#STU00212457).

## Results

Analysis of spiked DBS samples indicated a strong dose-response between CR3022 concentration and OD in wells coated with RBD at 2.0 μg/mL for both protocols (Figure 1). There was no response in wells with no capture antigen, although overall OD levels were higher for Protocol 1 than Protocol 2. The responses were approximately linear up to 6.25 μg/mL, with a flattening of response at higher concentrations of CR3022.

**Figure 1.**
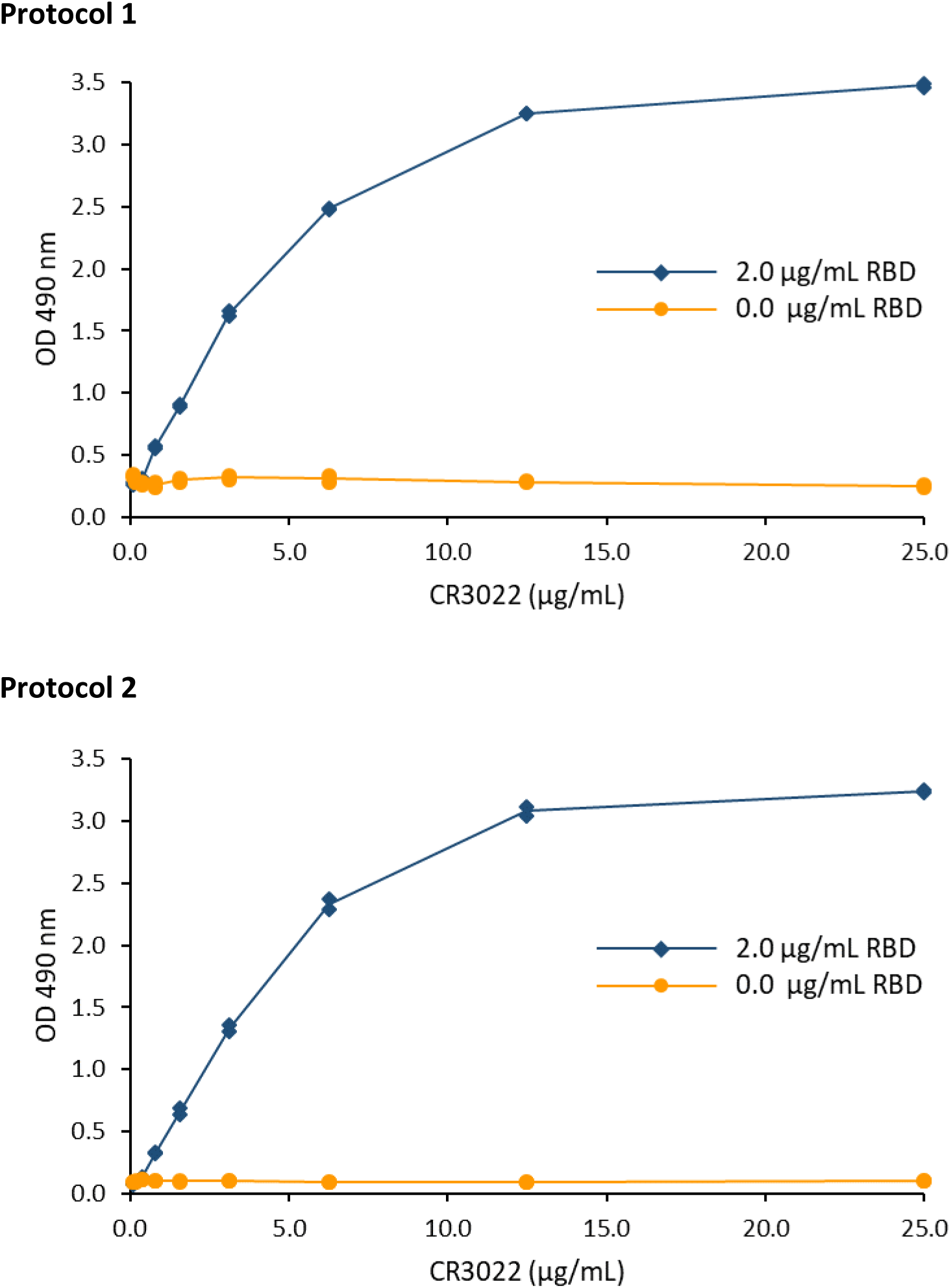
OD response to DBS samples spiked with known concentrations of CR3022 antibody in wells coated with RBD antigen in comparison with uncoated wells. Results for Protocol 1 are on top, Protocol 2 is below.

For Protocol 1, responses were significantly higher in DBS samples from PCR positive confirmed cases in comparison with presumptive negatives (one-way t = 1.99, p=0.04) (Figure 2). For positives, OD values ranged from 0.43 to 3.58, with a mean of 1.38. For negative samples, OD values ranged from 0.18 to 0.25, with a mean of 0.21 (SD=0.028).

**Figure 2.**
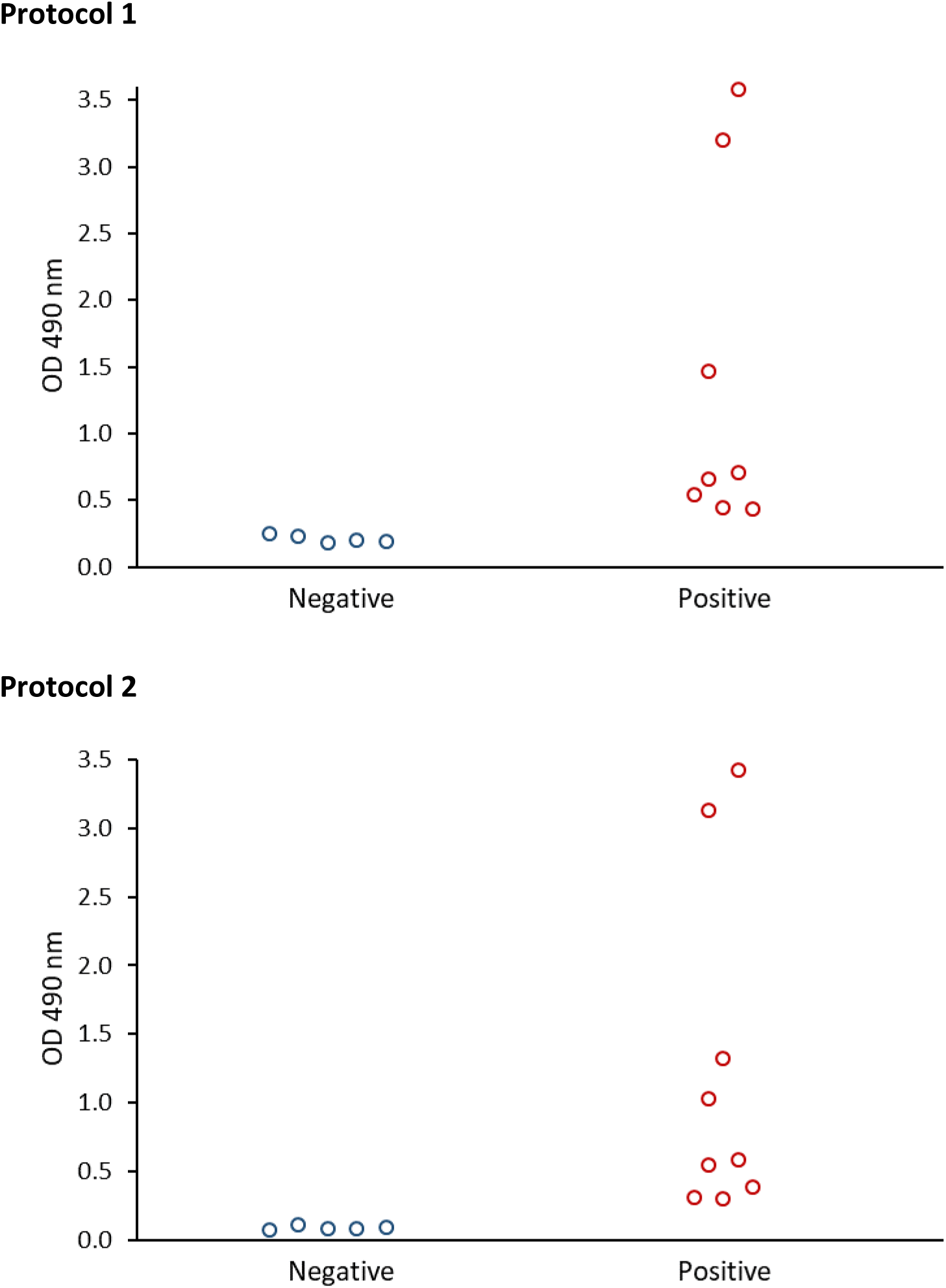
OD responses for DBS samples from confirmed PCR positive cases in comparison with presumptive negatives. Differences between positives and negatives are statistically significant for Protocol 1 (top; one-way t = 1.99, p=0.04) and Protocol 2 (below; one-way t = 2.05, p=0.03). One positive case did not have sufficient DBS material for analysis with Protocol 1.

Similarly, PCR positive cases generated significantly higher responses than negatives in Protocol 2 (one-way t = 2.05, p=0.03). For positives, OD values ranged from 0.31 to 3.41, with a mean of 1.22. For negative samples, OD values ranged from 0.071 to 0.11, with a mean of 0.09 (SD=0.014). Even though responses were shifted upward in Protocol 1, the level of agreement in results across protocols was very high: The Pearson correlation in OD values was 0.999 (p<0.0001).

## Discussion

COVID-19 is a deadly clinical disease that is now widely acknowledged to have high rates of community spread, where it is having devastating social and economic impacts^17,18^. Strategic testing in community-based settings is therefore critical for tracking the spread of SARS-CoV-2, and for identifying the factors that mitigate transmission. We have developed two DBS-based ELISA protocols for SARS-CoV-2 IgG antibodies that require only a single drop of blood, collected from a simple finger stick.

Our results demonstrate the feasibility of using DBS samples for serological testing of SARS-CoV-2 IgG antibodies. Responses are significantly higher for PCR confirmed positive cases, with separation in OD values between positive and negative samples. Additional analyses with a larger set of samples, including samples from suspected but not confirmed cases, will be needed to determine the criteria that should be applied for differentiating seropositive from seronegative individuals using DBS ELISA. Dilutions in DBS of an antibody like CR3022, which has known affinity, may be useful in this regard, particularly given the strong dose-response across the measurement range and the potential for application as calibration material across plates.

Advantages of DBS include a low cost and non-invasive approach to blood collection, and unlike serum/plasma, DBS samples do not need to be centrifuged and separated prior to transport and analysis^10^. Antibodies remain stable in DBS for several weeks at room temperature, and requirements for shipping are minimal^19^. Samples can therefore be self-collected by individuals in the home, and sent to the lab through regular mail for analysis. While point-of-care lateral flow immunoassay tests share some of the advantages of DBS sampling, they are difficult to implement in the home and they do not provide the same level of accuracy and quantification as lab-based tests for SARS-CoV-2 antibodies^12^.

Widespread serological testing for SARS-CoV-2 antibodies is essential for determining the level and distribution of infection in the community. As the first waves of infection subside, these data are necessary for ascertaining the true seroprevalence and mortality rate of infection, and for empirical analyses of the policies and individual behaviors (e.g., closing schools and businesses, social distancing) that reduce transmission. In addition, research on virus neutralization is ongoing to determine the extent to which antibodies confer protection against SARS-CoV-2 infection/reinfection^20,21^, and estimates of immunity in the general population are critical for forecasting the course of future outbreaks, the demand on limited clinical resources, and eventually, plans for vaccine roll out.

Serological testing with large samples drawn from the general population, as well as strategic sampling of subgroups of particular interest, is needed to achieve these goals. The logistical constraints associated with venous blood collection will make it difficult to implement widespread testing exclusively with serum-based tests. We therefore developed a DBS-based serological test for SARS-CoV-2 IgG antibodies which combines the convenience of sample collection in the home with the quantitation of ELISA in the lab in order to facilitate testing of large numbers of people, across a wide range of geographic contexts.

## Data Availability

Data to be uploaded as supplementary material.

## Acknowledgments

This research was supported with funding from the Canadian Institute for Advanced Research and Northwestern University.

